# Naïve Bayes is an interpretable and predictive machine learning algorithm in predicting osteoporotic hip fracture in-hospital mortality compared to other machine learning algorithms

**DOI:** 10.1101/2024.05.10.24307161

**Authors:** Jo-Wai Douglas Wang

## Abstract

Osteoporotic hip fractures (HFs) in the elderly are a pertinent issue in healthcare, particularly in developed countries such as Australia. Estimating prognosis following admission remains a key challenge. Current predictive tools require numerous patient input features including those unavailable early in admission. Moreover, attempts to explain machine learning [ML]-based predictions are lacking. We developed 7 ML prognostication models to predict in-hospital mortality following minimal trauma HF in those aged ≥ 65 years of age, requiring only sociodemographic and comorbidity data as input. Hyperparameter tuning was performed via fractional factorial design of experiments combined with grid search; models were evaluated with 5-fold cross-validation and area under the receiver operating characteristic curve (AUROC). For explainability, ML models were directly interpreted as well as analyzed with SHAP values. Top performing models were random forests, naïve Bayes [NB], extreme gradient boosting, and logistic regression (AUROCs ranging 0.682 – 0.696, p>0.05). Interpretation of models found the most important features were chronic kidney disease, cardiovascular comorbidities and markers of bone metabolism; NB also offers direct intuitive interpretation. Overall, we conclude that NB has much potential as an algorithm, due to its simplicity and interpretability whilst maintaining competitive predictive performance.

**Author Summary:** Osteoporotic hip fractures are a critical health issue in developed countries. Preventative measures have ameliorated this issue somewhat, but the problem is expected to remain in main due to the aging population. Moreover, the mortality rate of patients in-hospital remains unacceptably high, with estimates ranging from 5-10%. Thus, a risk stratification tool would play a critical in optimizing care by facilitating the identification of the susceptible elderly in the community for prevention measures and the prioritisation of such patients early during their hospital admission. Unfortunately, such a tool has thus far remained elusive, despite forays into relatively exotic algorithms in machine learning. There are three major drawbacks (1) most tools all rely on information typically unavailable in the community and early during admission (for example, intra-operative data), limiting their potential use in practice, (2) few studies compare their trained models with other potential algorithms and (3) machine learning models are commonly cited as being ‘black boxes’ and uninterpretable. Here we show that a Naïve Bayes model, trained using only sociodemographic and comorbidity data of patients, performs on par with the more popular methods lauded in literature. The model is interpretable through direct analysis; the comorbidities of chronic kidney disease, cardiovascular, and bone metabolism were identified as being important features contributing to the likelihood of deaths. We also showcase an algorithm-agnostic approach to machine learning model interpretation. Our study shows the potential for Naïve Bayes in predicting elderly patients at risk of death during an admission for hip fracture.

## 1. Introduction

The osteoporotic hip fracture (HF) is a global issue with an estimated financial burden of 17 billion USD for the United States in 2002 and projected burden of £3.62 in 2023 for the United Kingdom [1, 2]. Estimates for short-term (in-hospital) mortality following HF have been placed in the vicinity of 2 – 10%, with an estimated mortality rate of 2.7% for HF hospitalisation in Australia [3–5]. In developed countries, though preventative measures (targeting reduction of hip fracture risk factors such as osteoporosis and falls) have reduced the age-standardized incidence rate of hip fractures, the absolute rate is increasing due to the ageing population [6]. In Australia, for instance, hospitalisations for HF in the elderly increased by almost 20% between 2006-07 and 2015-16 from 15 900 to 18 700 respectively [5]. With the trend towards an aged population expected to continue, including in Australia, HFs in the elderly will remain a relevant, and increasingly pressing challenge in healthcare.

One key aspect in HF assessment and management challenge is the prognostication of poor short-term outcomes. There exists a substantial amount of analysis from traditional statistical methods (such as logistical regression, LR) in identifying key risk factors for predicting poor outcomes, notably mortality, following HF and scoring tools that have risen to prominence are the Nottingham Hip Fracture Score (NHFS) and the orthopaedic-Physiological and Operative Severity Score for the enUmeration of Mortality and Morbidity (O-POSSUM) [7–10]. Most of these tools require a combination of both clinical, laboratory and intra-operative data; and the lack of laboratory and intra-operative data early during admission limits the use of such tools in early risk stratification.

Non-traditional mathematical algorithms, especially those associated with artificial intelligence (AI) and machine learning (ML), have become increasingly utilized in healthcare. A variety of ML algorithms, including regression-based methods, decision-tree based methods (i.e. decision trees [DT], Random Forests [RF], eXtreme Gradient Boosting [XGB] implementation), neural networks (NN), Naïve Bayes and support vector machines (SVM) have been used in the prognostication of patients in the general peri-operative [11–16] and peri-HF [17–20] period with varying degrees of success. However, most of these tools require data that is not readily available on admission (such as intra-operative data and laboratory data), much like the tools developed from traditional statistical methods and most do not predict short-term in-hospital mortality following HF.

Moreover, few studies compared multiple machine learning algorithms of different classes with each other. An exception to this was work performed by Forssten et al., who trained SVM, NB, and as well as LR (to be used as a baseline) models to predict the 1-year mortality post-HF, and found that LR outperformed all other models [17]. To our knowledge no study has trained a wider array of machine learning algorithms for prediction of short-term outcome following HFs. Tree-based methods have received the majority of attention. Another algorithm that has remarkable potential is naïve Bayes which is based on Bayes theorem, with the additional ‘naïve’ assumption that features are conditionally independent. It has been applied successfully across a wide variety of tasks in natural language processing (e.g. detection of spam email [21], text sentiment analysis, text/document classification) as well as in the medical field (e.g. the prognostication in cirrhotic patients following TIPS [22], prediction of 30-day mortality following HF [23], prediction of osteonecrosis of femoral head with cannulated screw fixation [24] and prediction of mortality in post-surgical intensive care unit patients [25]).

While predictive ability is an important characteristic of any prognostic tool, it is increasingly recognized that a desirable attribute of machine learning algorithm is that they are interpretable (or ‘explainable’) especially as ML models become increasingly complex [26, 27]. Recognition of this issue has led to the development of the subfield of ‘interpretable’ ML and, in particular, the development and application of the SHapley Additive exPlanations (SHAP), an approach based on cooperative game theory [28–33].

Our goal, was to train multiple machine learning models, specifically Bernoulli Naïve Bayes (NB), DT, RF, XGB, SVM, logistic regression (LR) and the multi-layer perceptron (MLP, a 3-layer NN) to predict in-hospital mortality for the elderly admitted with HF. We focus on using only those patient features that are readily available in the early phases during a hospital admission, i.e. sociodemographic and comorbidity data. The performances of each model would be compared to identify the most predictive algorithm. Finally, each predictive tool would be analyzed via direct interpretation of model and with calculation of SHAP values.

## 2. Results

### 2.1. Patient cohort characteristics

Of the 3625 patients in the cohort, age was distributed non-normally with median age of 84 (interquartile range of 10 years) and 2730 (75.3%) were female; 189 (5.2%) had in-hospital mortality. The most common comorbidity was hypertension (at 2045 [56.4%]). Details are present in Table 1 (with abbreviations defined below).

**Table 1.** Sociodemographic features, outcomes of HF cohort.

| Variable | Total Cohort<br>(N=3625) | Female<br>(N=2730, 75.31%) | Male<br>(N=895, 24.69%) | p value<sup>(1)</sup> |
| --- | --- | --- | --- | --- |
| **Sociodemographic features** |  |  |  |  |
| **Age (median [IQR])** | 84 [10] | 85 [10] | 82 [12] | <0.001 |
| **Aged > 80 years (n,%)** | 2457, 67.8% | 1937, 71.0% | 520, 58.1% | <0.001 |
| **PRCF resident (n,%)** | 1208, 33.3% | 950, 34.8% | 258, 28.8% | 0.001 |
| **Smoker (n,%)** | 180, 5.0% | 124, 4.5% | 56, 6.3% | 0.050 |
| **Alcohol overuse (n,%)<sup>(2)</sup>** | 144, 4.0% | 60, 2.2% | 84, 9.4% | <0.001 |
| **Walking aids user (n,%)** | 1300, 35.9% | 999, 36.6% | 301, 33.6% | 0.116 |
| **Comorbidities features** |  |  |  |  |
| **Hypertension, (n,%)** | 2045, 56.4% | 1606, 58.8% | 439, 49.1% | <0.001 |
| **Anaemia (n,%)** | 1531, 42.2% | 1051, 38.5% | 480, 53.6% | <0.001 |
| **CKD (n,%)** | 1444, 39.9% | 1106, 40.5% | 338, 37.8% | 0.152 |
| **Dementia (n,%)** | 1117, 30.8% | 858, 31.4% | 259, 28.9% | 0.172 |
| **CAD (n,%)** | 1073, 29.6% | 750, 27.5% | 323, 36.1% | <0.001 |
| **History of AMI (n,%)** | 287, 7.9% | 191, 7.0% | 96, 10.7% | <0.001 |
| **AF (n,%)** | 702, 19.4% | 513, 18.8% | 189, 21.1% | 0.139 |
| **COPD (n,%)** | 561, 15.5% | 385, 14.1% | 176, 19.7% | <0.001 |
| **T2DM (n,%)** | 482, 13.3% | 325, 11.9% | 157, 17.5% | <0.001 |
| **OP (n,%)** | 478, 13.2% | 410, 15.0% | 68, 7.6% | <0.001 |
| **CVA (n,%)** | 431, 11.9% | 323, 11.8% | 108, 12.1% | 0.897 |
| **TIA (n,%)** | 309, 8.5% | 227, 8.3% | 82, 9.2% | 0.474 |
| **PD (n,%)** | 172, 4.7% | 97, 3.6% | 75, 8.4% | <0.001 |
| **Malignancy (n,%)** | 82, 2.3% | 52, 1.9% | 30, 3.4% | 0.017 |
| **PTH>6.8pmol/L** | 1684, 46.5% | 1275, 46.7% | 409, 45.7% | 0.628 |
| **25(OH)vitamin D≤25nmol/L** | 610, 16.8% | 467, 17.1% | 143, 16.0% | 0.464 |
| **25(OH)vitamin D≤50nmol/L** | 1659, 45.8% | 1235, 45.2% | 424, 47.4% | 0.283 |
| **Outcome** |  |  |  |  |
| **Died (n,%)** | 189, 5.2% | 130, 4.8% | 59, 6.6% | 0.040 |

<sup>1</sup>Pearson's Chi-squaerr test (Yates corrected).
<sup>2</sup>Use>3 times a week.
**Abbreviations:** PRCF, permanent residential care facility; CKD, chronic kidney disease; CAD, coronary artery disease; AMI, acute myocardial infarction; AF, atrial fibrillation; COPD, chronic obstructive pulmonary disease; T2DM, type 2 diabetes mellitus; OP, osteoporosis; CVA, cerebrovascular accident; TIA, transient ischaemic attack; PD, Parkinson's disease; PTH, parathyroid hormone

### 2.2. Model Performance – Training

The model with the highest area under the receiver operating characteristic (AUROC) was MLP (AUROC 0.828) followed by LR, RF, XGB and NB (0.733, 0.730, 0.726 and 0.725 respectively, all p>0.05), then DT (AUROC of 0.697) and finally SVM (AUROC 0.533).

The model with greatest area under the precision-recall curve (AUPRC) was MLP (AUPRC 0.245), followed by LR, XGB and RF (AUPRCs of 0.134, 0.133 and 0.130 respectively, p>0.05), NB (AUPRC 0.124), DT (AUPRC of 0.094) and finally SVM (AUPRC of 0.058). Details are present in Table 2 and Table 3.

**Table 2.** Model performance (training phase)

|  | AUROC |  |  | AUPRC |  |  |
| --- | --- | --- | --- | --- | --- | --- |
|  | Mean | STD | 95%CI | Mean | STD | 95%CI |
| **SVM** | 0.533 | 0.029 | 0.475 - 0.591 | 0.058 | 0.004 | 0.050 - 0.067 |
| **NB** | 0.725 | 0.007 | 0.711 - 0.739 | 0.124 | 0.003 | 0.117 - 0.131 |
| **LR** | 0.733 | 0.008 | 0.717 - 0.750 | 0.134 | 0.004 | 0.127 - 0.141 |
| **DT** | 0.697 | 0.004 | 0.690 - 0.704 | 0.094 | 0.001 | 0.093 - 0.095 |
| **RF** | 0.730 | 0.007 | 0.716 - 0.745 | 0.130 | 0.003 | 0.125 - 0.136 |
| **XGB** | 0.726 | 0.007 | 0.711 - 0.741 | 0.133 | 0.005 | 0.122 - 0.144 |
| **MLP** | 0.828 | 0.008 | 0.813 - 0.844 | 0.245 | 0.030 | 0.186 - 0.305 |

**Table 3.**
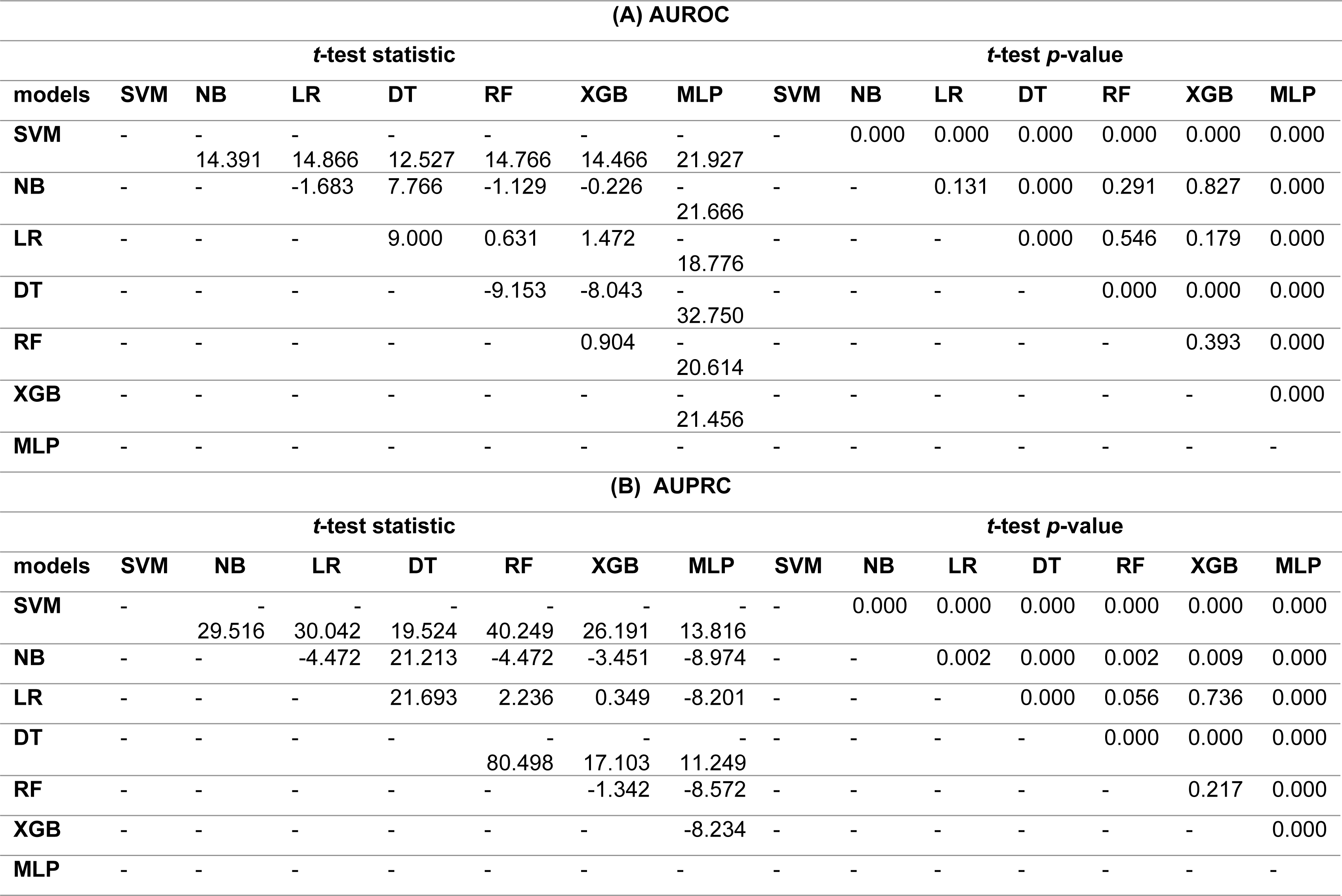
Comparison of model performance during training. (A) – AUROC (B) – AUPRC.

### 2.3. Model Performance – Test

The models with highest AUROC were RF, NB, XGB and LR (AUROCs of 0.696, 0.694, 0.689 and 0.682 respectively, all p>0.05;) followed by MLP and DT (AUROCs of 0.618, 0.616 respectively, p>0.05) and finally SVM (AUROC of 0.499).

The model(s) with highest AUPRC were NB, XGB, RF and LR (AUPRCs of 0.113, 0.113, 0.112, 0.112 respectively, p>0.05), MLP and DT (AUPRCs of 0.077, 0.070 respectively, p>0.05), and SVM (AUPRC of 0.054) – see Table 4, Table 5 and Figure 1.

**Figure.1.**
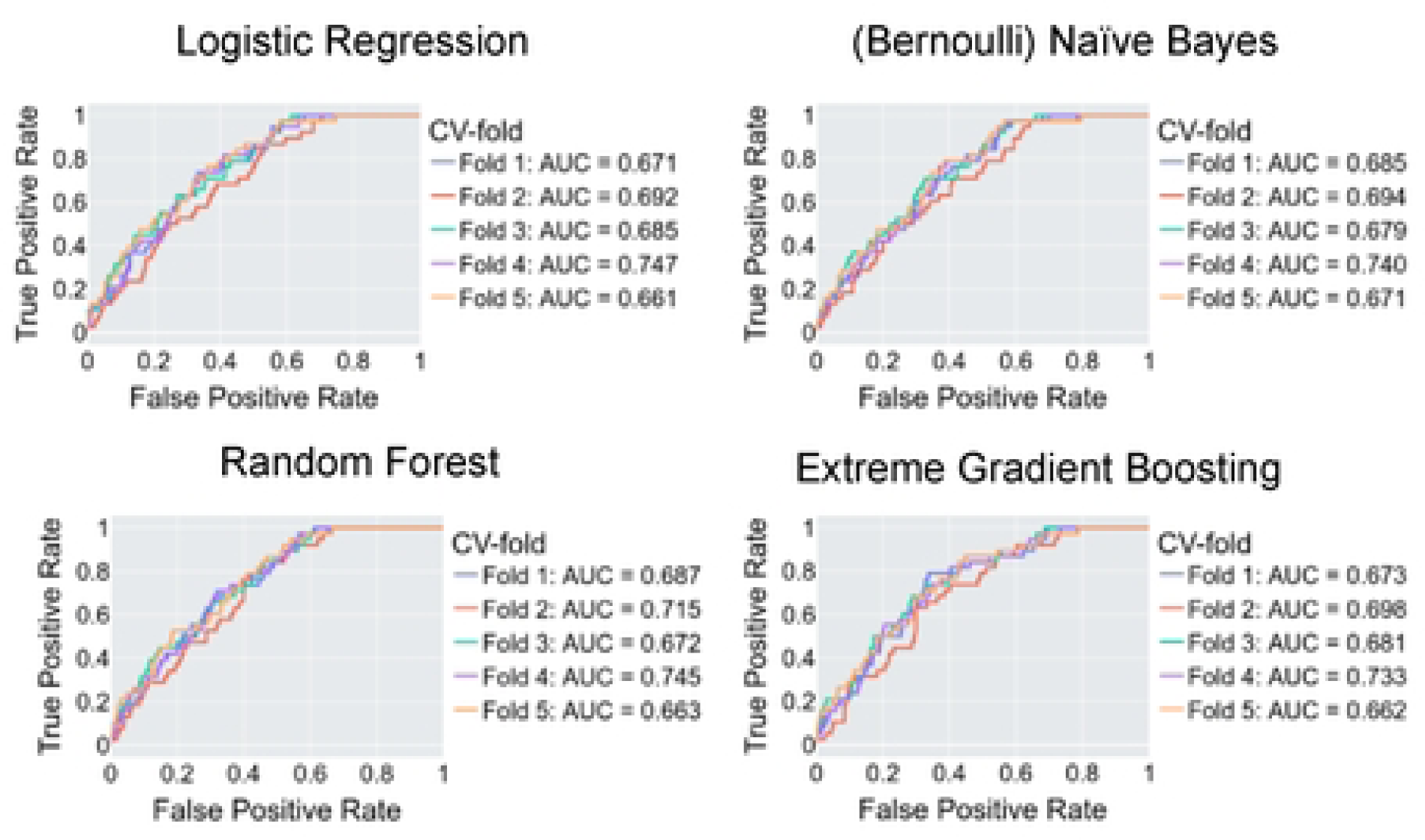
ML model test performance (area under the receiver operating characteristic. AUC). Only the test set AUCs evaluated from the o-fold cross-validation for the fourbesf-performing ML models are shown.

**Table 4.** Model performance during testing (5-fold cross-validation)

|  | AUROC |  |  | AUPRC - Average precision |  |  |
| --- | --- | --- | --- | --- | --- | --- |
|  | Mean | STD | 95%CI | Mean | STD | 95%CI |
| **SVM** | 0.499 | 0.048 | 0.404 - 0.594 | 0.054 | 0.006 | 0.041 - 0.067 |
| **NB** | 0.694 | 0.024 | 0.646 - 0.742 | 0.113 | 0.019 | 0.076 - 0.151 |
| **LR** | 0.682 | 0.034 | 0.614 - 0.750 | 0.112 | 0.019 | 0.074 - 0.150 |
| **DT** | 0.616 | 0.028 | 0.560 - 0.671 | 0.070 | 0.006 | 0.058 - 0.081 |
| **RF** | 0.696 | 0.030 | 0.636 - 0.756 | 0.112 | 0.024 | 0.064 - 0.161 |
| **XGB** | 0.689 | 0.025 | 0.640 - 0.738 | 0.113 | 0.024 | 0.065 - 0.160 |
| **MLP** | 0.618 | 0.027 | 0.565 - 0.671 | 0.077 | 0.007 | 0.063 - 0.090 |

**Table 5.**
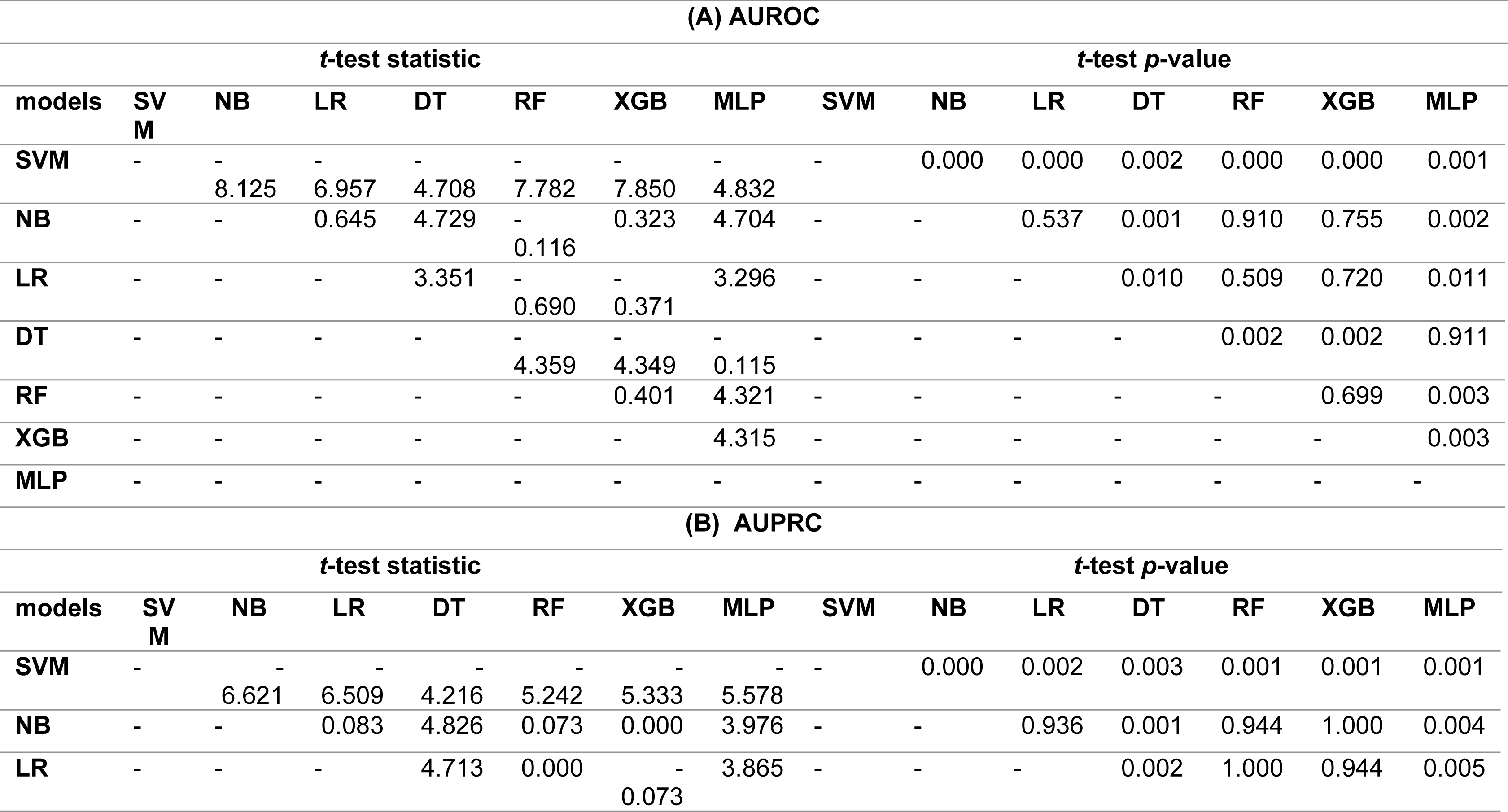
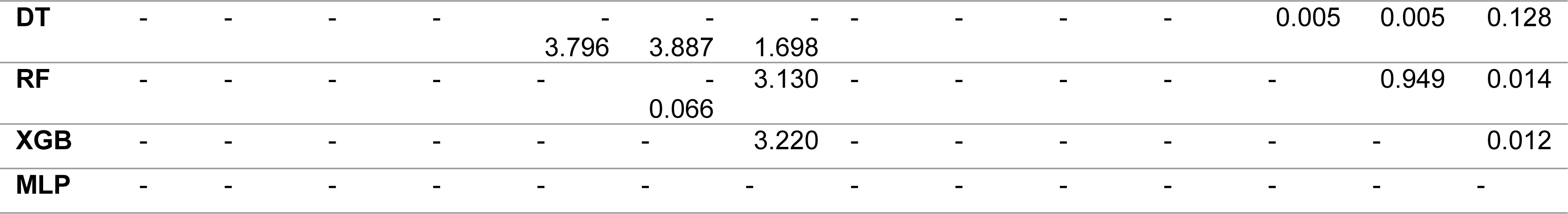
Comparison of model performance on testing. (A) – AUROC (B) – AUPRC.

### 2.4. Feature importance – Model interpretation

Feature importance rankings (1 being the most important) according to each model can be found in Table 6. Corresponding coefficients for NB, LR, XGB and RF can be found in Table 7.

**Table 6.** Feature importance rankings.

| Feature | Feature importance rankings* |  |  |  |  |  |  |
| --- | --- | --- | --- | --- | --- | --- | --- |
|  | LR | SVM | NB | DT | RF | XGB | MLP |
| **Male** | 9 | 6 | 13 | 12 | 12 | 12 | 3 |
| **Aged > 80** | 3 | 1 | 12 | 5 | 5 | 3 | 14 |
| **Resident of PRCF** | 10 | 20 | 7 | 7 | 7 | 6 | 13 |
| **Smoking** | 13 | 17 | 20 | 15 | 15 | 19 | 23 |
| **Alcohol overuse** | 22 | 22 | 23 | 16 | 16 | 20 | 21 |
| **Walking aids use** | 16 | 5 | 14 | 17 | 17 | 13 | 6 |
| **HT** | 18 | 7 | 19 | 18 | 18 | 22 | 20 |
| **CAD** | 8 | 14 | 4 | 3 | 3 | 2 | 19 |
| **MI** | 7 | 8 | 1 | 8 | 8 | 7 | 5 |
| **AF** | 5 | 15 | 2 | 13 | 13 | 5 | 1 |
| **CVA** | 20 | 11 | 16 | 19 | 19 | 16 | 22 |
| **TIA** | 15 | 13 | 11 | 20 | 20 | 15 | 10 |
| **dementia** | 12 | 10 | 8 | 4 | 4 | 9 | 4 |
| **PD** | 14 | 19 | 18 | 21 | 21 | 17 | 18 |
| **COPD** | 4 | 12 | 6 | 11 | 11 | 11 | 12 |
| **T2DM** | 17 | 16 | 15 | 10 | 10 | 14 | 11 |
| **CKD** | 1 | 2 | 3 | 1 | 1 | 1 | 2 |
| **Anaemia** | 11 | 4 | 10 | 9 | 9 | 10 | 8 |
| **Malignancy** | 23 | 23 | 22 | 22 | 22 | 23 | 17 |
| **Osteoporosis** | 21 | 21 | 21 | 23 | 23 | 18 | 16 |
| **PTH>6.8pmol/L** | 6 | 18 | 9 | 2 | 2 | 4 | 7 |
| **25(OH)vitamin D≤25nmol/L** | 2 | 9 | 5 | 6 | 6 | 8 | 9 |
| **25(OH)vitamin D≤50nmol/L** | 19 | 3 | 17 | 14 | 14 | 21 | 15 |

\*In descending order of importance.

For the LR model, the 5 most important patient features in prediction of mortality were presence of CKD, vitamin D deficiency (≤25nmol/L), advanced age (>80 years), COPD, and AF. In the SVM model the 5 most important patients features in prediction of mortality were advanced age (>80 years), CKD, vitamin D insufficiency (≤50nmol/L), anaemia, and use of walking aids. For the NB model, the 5 most important features in mortality prediction were history of MI, AF, CKD and CAD. For the DT model the 5 most important features in mortality prediction were presence of CKD, hyperparathyroidism (PTH>6.8pmol/L), CAD, dementia and advanced age (>80 years). For the RF model the 5 most important features in mortality prediction were CKD, hyperparathyroidism (PTH>6.8pmol/L), CAD, dementia and advanced age (>80 years). For the XGB model the 5 most important features in mortality prediction were CKD, CAD, advanced age (>80 years), PTH>6.8pmol/L and AF. Finally, for the MLP model the 5 most important features in mortality prediction were AF, CKD, male sex, dementia, and MI.

### 2.5. Feature importance – SHAP analysis

Features were also ranked by the mean absolute SHAP values as displayed in summary below (Figure 2).

**Figure 2.**
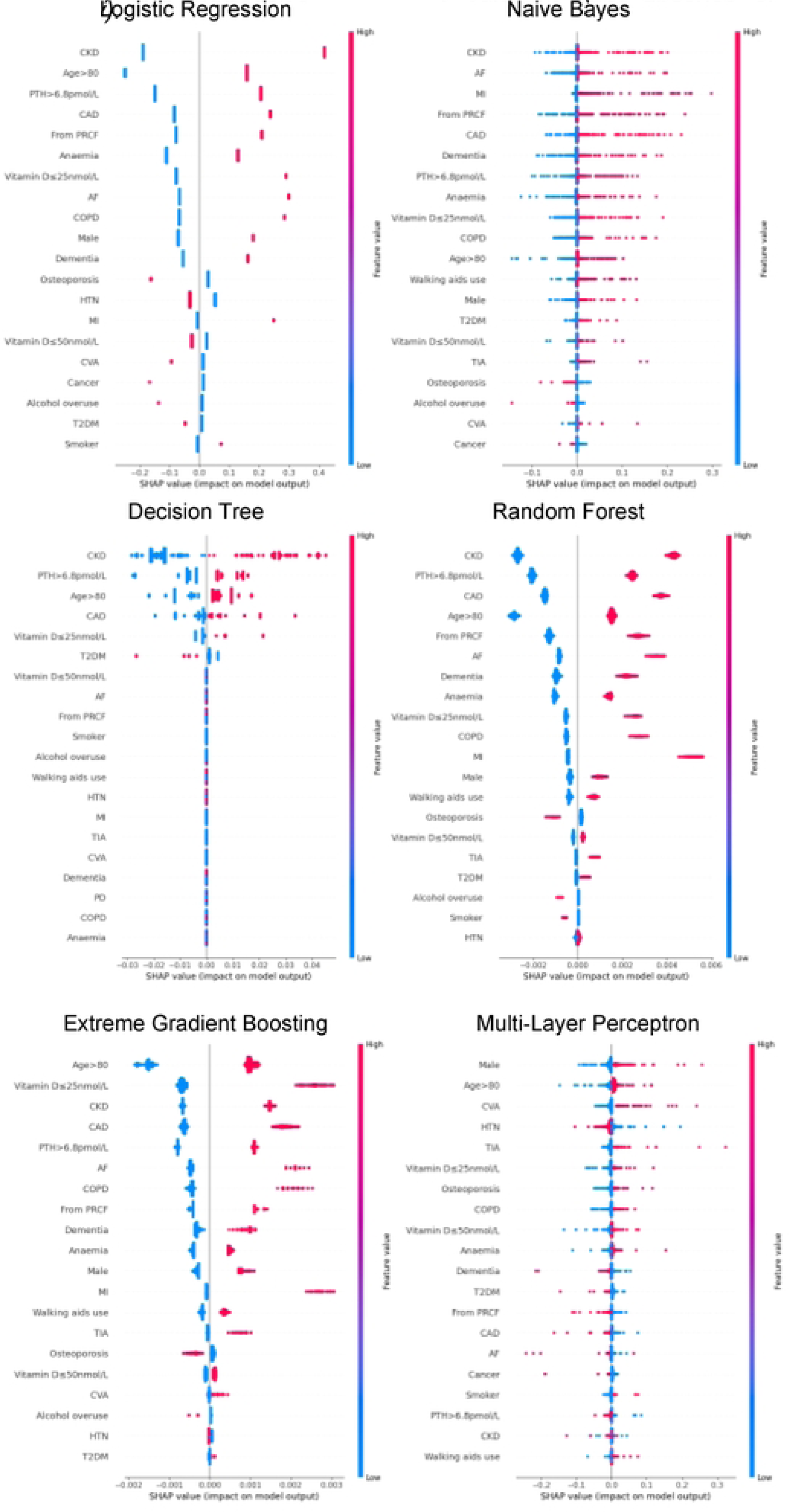
**Summary plot SHAP values for patient comorbidities.** Each point on the plot represents a SHAP value for an individual patient’s comorbidity ($HAP value is on x-axis, corresponding comorbiditv on the y-axis. Positive SHAP values corresponas to a positive/additive contribution to the Qrediction (i.e. in-hospital mortality); conversely a negative SHAP value corresponds to a negative/subtractive contribution. Colours of points represents feature varues: maintalred corresponded to *a* value of ‘1’ **(i.e.** presence of the *comorbidi and blue corresponding to value ‘O’ (i.e. absence of comorbidi* gistic Regression

For the LR model, the 5 most predictive patient features for mortality in order from highest magnitude to lowest, based on mean SHAP values, were CKD, advanced age (>80 years), hyperparathyroidism (PTH>6.8pmol/L), CAD, and residency from PRCF. Absence of any of these features had a negative SHAP value (i.e. a negative contribution) on the model outcome (in-hospital mortality); the magnitude of this impact was consistent across all patients. Likewise, the presence of any of these features always had a positive SHAP value (i.e. an additive contribution) on in-hospital mortality. The magnitude of this effect was again consistent across all patients.

For NB model, the 5 most predictive features were CKD, AF, MI, residency from PRCF, CAD. Again, absence of any of these features most commonly had a negative impact on in-hospital mortality; the magnitude of this effect varied among patients. The presence of any of the above 5 features had a positive contribution to the prediction of in-hospital mortality; similarly, the magnitude of this effect varied significantly among patients.

For the DT model, the 5 most predictive features were CKD, hyperparathyroidism (PTH>6.8pmol/L), advanced age (>80 years), presence of CAD and vitamin D deficiency (≤25nmol/L). Presence of these five comorbidities had a positive contribution to prediction of in-hospital mortality and, conversely their absence had a negative contributory effect on prediction. Interestingly, absence of T2DM had an additive effect and presence of T2DM had a negative effect on mortality prediction. The magnitude of contributions that each of the 5 variables had varied among different patients. Finally, it is noteworthy that all other comorbidities had little to no influence on patient outcomes.

For the RF model, the 5 most predictive features were CKD, hyperparathyroidism (PTH>6.8pmol/L), CAD, advanced age (>80 years) and residence from PRCF. The presence of these features increased likelihood of mortality and conversely absence decreased the likelihood of mortality; there was only a minor variation of contribution from each feature for each patient.

For the XGB model, the 5 most predictive patient features were advanced age (>80 years), vitamin D deficiency (≤25nmol/L), CKD, CAD and hyperparathyroidism (PTH>6.8pmol/L). The presence (and absence) of any of these features increased (or decreased) the likelihood of mortality. For each feature, there was only mild variation in the magnitude of contributions among patients.

Finally, from the MLP model, the 5 most predictive patient features were male sex, advanced age (>80 years), CVA, HTN and TIA. The presence (or, conversely, the absence) of any of these features except for HTN were associated with an increased (decreased) likelihood of mortality; presence (or absence) of HTN appeared to decrease (increase) the likelihood of mortality.

Across all models, the 5 comorbidities most consistently with the greatest influence on mortality prediction were: CKD, advanced age (>80 years), elevated PTH (>6.8pmol/L), cardiovascular disease (CAD, MI, AF or HTN) and PRCF residence.

## 3. Discussion

We have trained and compared the performances of 7 machine learning models to predict in-hospital mortality for hospitalized elderly minimal trauma HF patients using only categorical data. Overall, the models had reasonable to good performance. We also performed an analysis of each model and applied SHAP analysis to gain insight into feature importance.

### 3.1. Model performance – Training and Test

Notably, but unsurprisingly, classification performance showed some variation among algorithms. The trained models, ordered in decreasing performance (based on both test AUROCs and test AUPRCs), were RF, NB, XGB and LR (all with no statistically significant difference in performance – see Table 4, 5 and Figure 1) followed by MLP and DT (no statistically significant difference in performance) and finally SVM. AUROCs ranged from 0.500 (SVM) to almost 0.700 (good performance), while AUPRC values ranged from 0.050 (SVM) to 0.115; a reflection of using a simplified model (with binary input data) to perform predictions on a minority class in this imbalanced dataset. There was minimal difference between the training and cross-validation performance for the top 4 models (RF, NB, XGB and LR). A greater variation in training and cross-validation performance scores was noted for DT and MLP, an indicator of overtraining (an infamous tendency in machine learning). That overtraining has occurred despite systematic and meticulous hyperparameter tuning, is strongly suggestive of insufficient data.

To our knowledge, most studies have focused on only training and applying one class of machine learning algorithm. Often there is no baseline model trained using traditional statistics (e.g. LR). Indeed, most studies have solely utilized tree-based methods (e.g. applying DT, XGB and RF methods) and this is reflected in a scoping study of ML usage in health economics and research (on 805 studies) which found the most frequent algorithms used were tree-based methods followed by regression-based (linear/logistic) methods, SVM, NN and finally NB [37]. However, it is known that performance on various tasks varies with different ML algorithms [38] and our finding that predictive performance varies among machine learning algorithms (for the same problem, using the same data) is consistent with this. It is thus ideal that in future applications of machine learning, a more comprehensive set of algorithms are trained, or some justification should be provided if possible when certain algorithms are not included.

The performance of NB in predicting mortality is on par with RF, XGB and LR which warrants further discussion here as it has received relatively little attention in the literature. Key to its success is the simplifying assumption of conditional independence among all patient input features. The most obvious advantage from this is that, by virtue of such a simplification, it is computationally inexpensive and is fast to train and run. However, with such a large seemingly excessive, simplifying assumption (not strictly satisfied in our current database), it may seem surprising that this model performs so well. Contrary to intuition, its good performance is not a coincidental or even unexpected phenomenon; formal analysis of NBs has established it performs well because the interdependencies, when they do exist, occur in a manner which results in them ‘cancel[ling] each other out’ [39].

### 3.2. Feature importance – Model interpretation and SHAP analysis

Rankings of patient comorbidity importance in their role in mortality prediction were determined from all models from direct interpretation of feature coefficients (see Table 6 and Table 7). CKD was most consistently ranked as one of the 5 most important patient comorbidities in predicting mortality. The other most important patient features included markers reflective of bone metabolism (PTH, vitamin D levels) and cardiovascular disease (presence of either one of CAD, MI, AF). Similar trends were found via SHAP value analyses for each model, i.e. CKD, bone metabolism markers and presence of cardiovascular diseases had the strongest influence on prediction of mortality based on mean SHAP values (Figure 2). It is recognized in the literature that cardiovascular comorbidities and renal function are important for prognostication which is reflected in their inclusion as input parameters for non-cardiac surgery risk assessment tools such as the Revised Cardiac Risk Index and the American College of Surgeons - surgical risk calculator [40–44]. However, these features are not explicitly included in HF-specific risk assessment tools (e.g. in O-POSSUM only symptoms and clinical findings suggestive of cardiovascular disease are included and NHFS only the number of comorbidities is included as an input parameter) [7–10]. Moreover, neither PTH or vitamin D levels are included in any of the current tools, despite an increasing number of studies supporting the key role they play in bone metabolism and prevention of fracture [45–57] and, potentially, with increasing recognition of their importance in the immunity [58–60] prevention of post-operative complications such as hospital acquired infections.

### 3.3. Further insights from model analysis

Of the four most predictive models, NB and LR models offer intuitive, quantifiable insights into feature contributions to prediction: in LR, the odds ratio can be taken by calculating the exponent of the coefficients, while in NB, from our method of scoring input features (see Appendix A), each coefficient corresponds to the ratio of the rate of the comorbidity in those who experienced in-hospital mortality compared to the comorbidity rate in those who survived. So, for example, in predicting mortality, we can see from the LR model that CAD, with a score of 0.319 (95%CI 0.180 – 0.458) increased mortality risk by 37% (OR 1.37; 95%CI 1.20 – 1.58) and CKD, with a score of 0.711 (95%CI 0.505 – 0.918) increased mortality risk by 2.03 (95%CI 1.66 – 2.50). From the NB model, a score of 1.62 (for CAD), and a score of 1.67 (for CKD) indicated that the rate of each comorbidity was greater in mortality than in survival by 62% and 67% respectively.

For the other top predictive models, insights gained from direct interpretation of RF and XGB is not so straightforward. Both these methods are based on DTs, which is itself an interpretable and intuitive model. However, a major drawback of DTs is that they are very prone to bias and variance (overfitting). RF and XGB address this issue by constructing multiple DTs and the overall prediction is then made from an ensemble/collection of multiple trees (numbering in the hundreds) and, hence, increased predictive performance is obtained at the expense of interpretability. In our study, the coefficients for each feature correspond to the relatively abstract concept of mean decrease in (Gini) impurity (see Appendix A).

### 3.4. Further insights from SHAP values

SHAP values revealed that the presence of more ‘severe’ comorbidities in each ML model had a more important additive effect on mortality risk than less severe comorbidities, as one might expect. For instance, patients with a history of acute MI (a higher severity subcohort of CAD patients) typically had the greatest SHAP value indicating that the presence of history of past MI had the greatest additive effect on mortality prediction. Similarly, the presence of vitamin D deficiency (≤25nmol/L) was correlated with greater SHAP values compared to vitamin D insufficiency (≤50nmol/L) (see Figure 2). In contrast, the absence of both MI and vitamin D deficiency in patients had less of a negative effect on mortality prediction compared to the other comorbidities (and hence was why they had a lower overall importance based on mean SHAP values). This reflects their relatively low prevalence in the cohort (Table 1).

For LR, RF, and XGB the SHAP values had low variability and were highly concentrated – an indication that the corresponding input patient features were consistently strong contributors to mortality prediction; a corollary of this was that these models offered good population level insight into mortality risk. Of the top four models, NB was the only model in which the SHAP values themselves varied among individuals. This variability in SHAP values among patients suggested that the influence of each singular comorbidity was not constant, and that each prediction appeared to be tailored toward individual.

### 3.5. Limitations

We note limitations to our study. Firstly, this was a study on a retrospective cohort, and all members of cohort were from a single-centre study. We recognize internal nested cross-validation, though relatively rigorous, is no substitute for external validation of our findings and that our tools need to be tested on external cohorts. Importantly, though the dataset used here is not of unreasonable size, we acknowledge that it may still be insufficient: firstly, because of the overfitting noted in MLP models and secondly because of the imbalance inherent to the issue class imbalance of mortality in HF – reflected in our dataset with a 5% mortality rate (and with only 191 cases the mortality population may be under-represented from a machine-learning perspective which typically requires cohort sizes numbering in the 1000s or greater to be trained effectively). We have restrained ourselves to conducting analysis using only categorical features. Model predictions were not calibrated, and it is known that certain machine learning models, particularly NB are notoriously poor at estimating probabilities despite being good classifiers.

### 3.6. Conclusion and final comments

In summary, NB was the most optimal ML model having the optimal virtues of strong predictive performance, model interpretability and potential for making individualized predictions. While RF, XGB and LR had similar performance capabilities, by nature they are not readily interpretable (i.e. RF and XGB) or are not optimal for individualized predictions (i.e. LR).

With ongoing development of digital infrastructure in the healthcare industry it is inevitable that machine learning algorithms will only become increasingly powerful and commonplace. As we await this reality, we hope that the findings here will provide physicians and clinicians with a tool that can be used to rapidly identify patients at higher risk of mortality early by knowledge of patient comorbidities; currently most prognostication tools can only be applied later in the admission. Moreover, we hope to provide valuable insights in applying machine learning models in healthcare for clinicians and researchers, in particular the advantages of the computationally inexpensive NB models highlighting its simplicity and interpretability with negligible compromise in performance.

## 4. Materials and Methods

### 4.1. Ethics Statement

The study was conducted in accordance with the Declaration of Helsinki (1964) and the Council for International Organisations of Medical Sciences International Ethic Guidelines and approved by the Australian Capital Territory Human Research Ethics Committee (reference number: 2023.LRE.00063). Because the analysis was based on a digital anonymized database, the patients’ written informed consent was waived.

### 4.2. Data Collection

Our cohort comprised 3625 elderly (i.e. aged ≥ 65 years of age) patients consecutively admitted to the Department of Orthopaedic Surgery at the Canberra Hospital between 1999 – 2019 with osteoporotic hip fracture. Patients admitted with hip fracture secondary to moderate-high energy trauma, or secondary to minimal trauma but with malignancy associated pathological fracture were excluded. Data on in-hospital mortality, sociodemographics (age, sex, smoking status, active history of overuse of alcohol, use of walking aids, and if the patient was a resident of an permanent residential care facility [PRCF]) and comorbidities (presence of hypertension [HT], coronary artery disease [CAD], previous history of acute myocardial infarction [MI], atrial fibrillation [AF], past history of stroke [cerebrovascular accident, CVA], transient ischaemic attack [TIA], dementia, Parkinson’s disease [PD], chronic obstructive pulmonary disease [COPD], type 2 diabetes mellitus [T2DM], chronic kidney disease [CKD], anaemia, history of solid organ malignancy, osteoporosis and hyperparathyroidism [parathyroid hormone/PTH>6.8pmol/L] and vitamin D insufficiency/deficiency; (25)OH vitamin D ≤ 50/25nmol/L) were collected.

### 4.3. Model Development

Seven machine learning algorithms, LR (as the baseline), SVM, NB, DT, RF, XGB, and the multi-layer perceptron (MLP, a 3-layer NN) were trained to predict mortality. For each algorithm the following steps were taken: identification of key hyperparameters to be trained (using a fractional factorial design of experiments approach), tuning of these key hyperparameters (using an inner 3-fold cross-validation to identify optimal hyperparameter and an outer 5-fold cross-validation to evaluate performance). Computations were performed using the Python packages, sklearn and pandas [34, 35].

### 4.4. Model Performance (and comparisons)

Performance was measured using the area under the receiver operating curve (AUROC) and the area under the precision-recall curve (AUPRC). Respective scores for each model were evaluated with 5-fold cross-validation; the mean and standard deviation of these scores was taken, and the 95% confidence interval was calculated. The student *t*-test was used to compare the mean performance scores. Computations were performed using the Python package SciPy (in particular ‘scipy.stats’ routines) [36].

### 4.5. Feature importance – Model Interpretation

Each trained model was analyzed directly. In general, the training of each model involved optimization of coefficients corresponding to each patient feature (comorbidity). The trained models were analyzed; for each patient comorbidity a corresponding coefficient or score was computed (see Appendix A). Features were ranked by importance based on the values of these scores.

### 4.6. Feature importance – SHAP analysis

For each patient, the SHAP value allocates a quantifiable credit to each variable (i.e. patient comorbidity) in its contribution to the model output (i.e. the final prediction). Feature importance analysis with SHAP was performed using the Python implementation [30, 33]. Features were ranked based on the mean SHAP values for each comorbidity.

## Data Availability

The raw data supporting the conclusions of this article will be made available by the authors on request, without undue reservation.

## 6. Appendix A

In the following section we give some mathematical context to the models used and show how the most predictive models (RF, XGB, NB and LR) were used to direct obtain feature importance coefficient/scores for each evaluation in the 5-fold cross validation. The average of these scores was used as the final feature score. The feature scores for NB and LR have the added benefit of offering intuitive insight as noted above in the discussion.

### 6.1. LR (logistic regression)

The LR predicts a probability using the function:

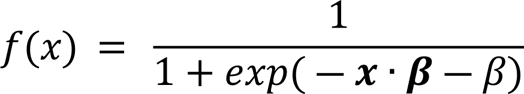

where *x* = [*x*_1_,*x*_2_,…,*x*_*n*_] is a vector for a patient encoding the presence or absence of a comorbidity (values of ‘1’ and ‘0’ respectively), **β** = [β_1_,β_2_,β_3_,…,β_n_] is a vector containing the coefficient weights (corresponding, in this study, to each patient comorbidity) and β the intercept value.

### 6.2. (Bernoulli) Naïve Bayes (NB)

Naïve Bayes uses the following classification rule:

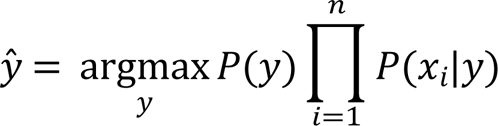

In Bernoulli Naïve Bayes (i.e. for binary classification, using binarized input):

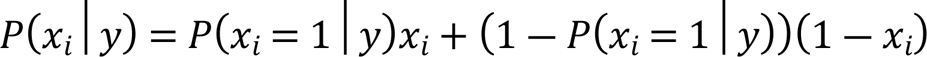

For each patient comorbidity *x*_*i*_, the model coefficient (i.e. the feature importance score) was computed as:

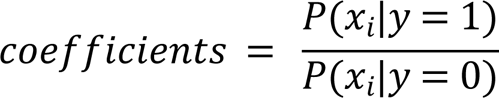

where *P*(*x*_*i*_|*y* = 1) and *P*(*x*_*i*_|*y* = 0) is the probability of the *i*^th^ comorbidity occurring given they experienced in-hospital mortality (*y* = 1) and survived to discharge (*y* = 0) respectively.

### 6.3. Tree-based methods (DT, RF, XGB)

In developing a single decision tree, the features are recursively partitioned to group patients by outcome (mortality). At each step (or ‘node’) the feature that results in the greatest reduction in ‘impurity’, or conversely the greatest increase in ‘purity’ is chosen. The measured used in this paper for all tree based methods (DT, XGB and RF) was the Gini impurity which is given by the formula:

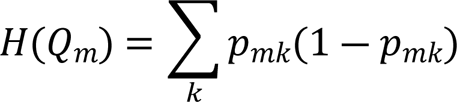

Where *p_mk_* is the proportion of those who died (k=1) or survived (k=0) at decision step (or node) number *m* and subsequently simplifies to:

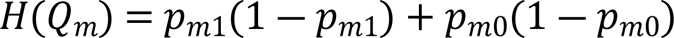

For RFs and XGBs where features may be used more than once (multiple trees are trained) the mean decrease in impurity across all nodes and trees is computed.

